# A multiplex serological assay for the characterization of IgG immune response to SARS-CoV-2

**DOI:** 10.1101/2021.09.23.21262329

**Authors:** Etienne Brochot, Vianney Souplet, Pauline Follet, Pauline Ponthieu, Christophe Olivier, Gaël Even, Christophe Audebert, Rémi Malbec

## Abstract

**Background:** In the fight against SARS-COV-2, the development of serological assays based on different antigenic domains represent a versatile tool to get a comprehensive picture of the immune response or differentiate infection from vaccination beyond simple diagnosis.

**Objectives:** Here we use a combination of the Nucleoprotein (NP), the Spike 1 (S1) and Spike 2 (S2) subunits, and the receptor binding domain (RBD) and N-terminal domain (NTD) of the Spike antigens from the Syrius-CoViDiag^®^ multiplex IgG assay, to follow the immune response to SARS-CoV-2 infection over a long time period and depending on disease severity.

**Results:** Using a panel of 209 sera collected from 61 patients up to eight months after infection, we observed that most patients develop an immune response against multiple viral epitope, but anti-S2 antibodies seemed to last longer. For all the tested IgGs, we have found higher titers for hospitalized patients than for non-hospitalized ones. Moreover the combination of the five different IgG titers increased the correlation to the neutralizing antibody titers than if considered individually.

**Conclusion:** Multiplex immunoassays have the potential to improve diagnostic performances, especially for ancient infection or mild form of the disease presenting weaker antibody titers. Also the combined detection of anti-NP and anti-Spike-derived domains can be useful to differentiate vaccination from viral infection and accurately assess the antibody potential to neutralize the virus.

## 1. Introduction

Since its first detection in Wuhan (China) in December 2019, the Severe Acute Respiratory Syndrome Coronavirus 2 (SARS-CoV-2) has rapidly spread to reach other countries worldwide as the coronavirus 2019 disease (COVID-19) became pandemic (1).

The virion has a nucleocapsid composed by genomic RNA and phosphorylated Nucleocapsid (NP) protein, which is buried inside a phospholipid bilayer and covered by the Spike proteins trimmers (S) that gives the CoVs their crown-like appearance on which their names are based. The S protein has two subunits, the Spike 1 (S1) which contains the receptor-binding domain (RBD) and N-terminal domain (NTD) and the Spike 2 (S2) (2). The choice of the antigenic domain is important, as it must be specific to the SARS-CoV-2 for discrimination against other hCoVs for example, and sensitive enough so infection would not be missed (Brochot et al., 2020). Also, anti-RBD antibodies are known to play a role in patients protection as this domain is used by the virus to penetrate host cells (4). Most commercial serological assays have demonstrated satisfying performances in terms of diagnostic sensitivity and specificity, based on one of those main different antigenic domains (5,6). However, the combination of different immunogenic antigens can give a more comprehensive picture of the humoral response strength and diversity (7–9) while maintaining elevated diagnostic performances (10,11). In multiplex assays, positivity thresholds can be adjusted to compensate for the use of antigenic domains more conserved between coronaviruses (12). Moreover, as vaccines are based on the Spike protein, the additional detection of anti-NP antibodies allows to differentiate viral infection from vaccination.

This study reports the design and use of the Syrius-CoViDiag^®^ multiplex IgG assay for the characterization of the immune response against over time, depending on disease severity, and in perspective of neutralizing antibody titers.

## 2. Material and Methods

### 2.1. Study design and cohort

The study was conducted at Amiens University medical Center (France). Samples were derived from de-identified excess serum specimens. The demographic information of the patients are available in Table 1. The study was approved by the institutional review board of the Amiens University Medical Center (number PI2020_843_0046, 21 April 2020).

**Table 1.** Cohort Characteristics.

|  |  |
| --- | --- |
| Number of patients | 61 |
| Female | 36 |
| Male | 25 |
| Age (Years):<br>Median<br>Range | 74<br>26-98 |
| >65 years | 41 |
| Hospitalized patients | 27 |
| Nonhospitalized patients | 34 |
| Immunocompromised patients | 6 (2 kidney transplant, 2 bone marrow transplant, 2 chemotherapy) |
| Number of samples (days post-PCR) |  |
| 0-59 | 52 |
| 60-119 | 49 |
| 120-179 | 49 |
| ≥180 | 59 |
| Number of patients (days post-PCR) |  |
| 0-59 | 50 |
| 60-119 | 36 |
| 120-179 | 42 |
| ≥180 | 46 |

Briefly, we used n=209 samples collected between March and April 2020 from n=61 patients (27 hospitalized patients and 34 non-hospitalized patients) with PCR-confirmed SARS-CoV-2 infections to perform immunoassay and virus seroneutralization test as already described in Aubry et al., 2021. All samples have been tested according to manufacturer’s instruction on the SirYus-CoViDiag® serological assay and the raw results are available in supplementary data.

### 2.2. Serological assay

The SirYus-CoViDiag® multiplex immunoassay targets IgG antibodies against five different antigens of the virus: NP, S1, S2, RBD, and NTD (Fig. 1). Note that the S1 and NP antigens have been printed in dot replicates in the shape of an “S” and “N” letters, respectively. This design allows for quick visual interpretation of seropositivity and vaccination status. The results have been automatically delivered using the SciReader® plate reader (Scenion GmbH) and associated analysis software, and an algorithm combining different cut-offs for the different antigens according to the manufacturer instructions. The spot mean signal intensity (MSI) was calculated as the average pixel value inside the spot perimeter minus the local background around the spot as described in Malbec et al., 2020. When multiplexing, the positivity thresholds can be adjusted with the number of different IgG antibodies detected. As NP and S2 antigens are more conserved between coronaviruses, SARS-CoV-2 IgG positivity is declared when a single one of them gives a signal over 40 MSI. However when NP and S2 antibodies are concomitantly present, the cut-off is adjusted to 20 MSI. For S1, RBD, and NTD antibodies a cut-off of 10 MSI is applied for SARS-CoV-2 positivity. Based on this algorithm, the test has been accredited by the French National Reference Center (CNR) in August 2020. On a reference cohort of 48 sera from patients positive to Covid-19 and 48 sera from patients negative to Covid-19. 14 days post symptoms, the diagnostic sensitivity was 90 %. The diagnostic specificity was 100 %.

**Figure 1.**
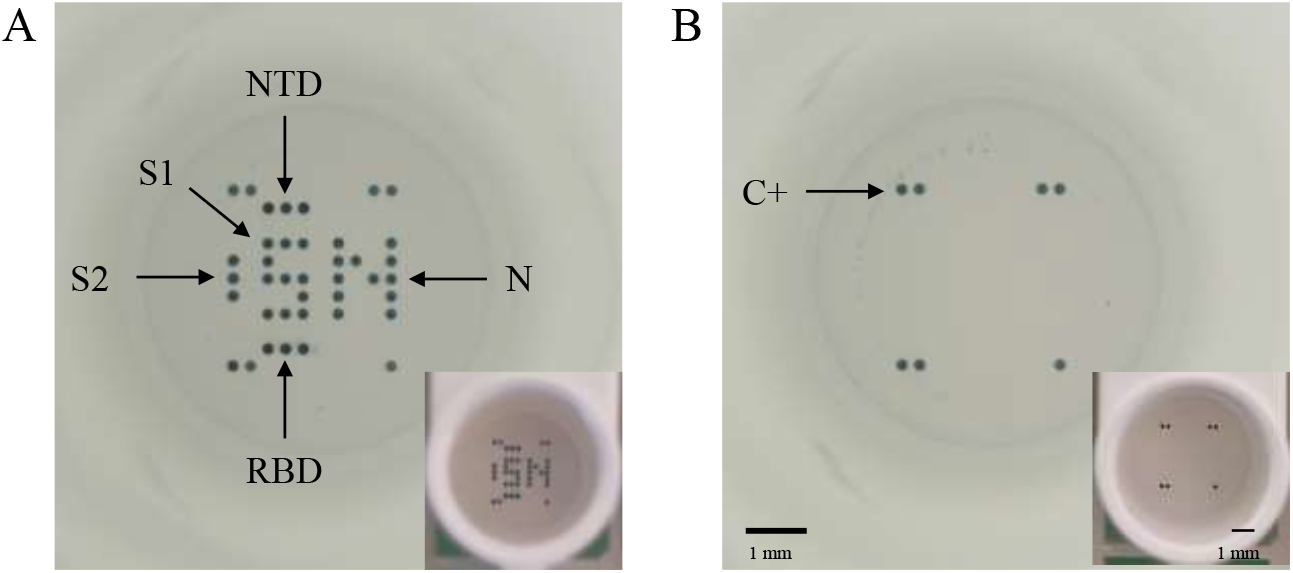
Full well pictures pictures obtained with the microplate reader (SciReader^®^) or with a phone camera (in insert) after incubation with the CoViDiag^®^ assay. (A) Positive sample presenting antibodies against the Nucleopcapside (NP), Spike 1 (S1), N-terminal domain (NTD) and Receptor binding domain (RBD) of the Spike protein, or Spike 2 (S2) antigens. (B) Negative sample with positive control on the edges. Scale bars correspond to 1 mm.

### 2.3. Statistical analysis

For the statistical analysis, Student’s test was used to test the relationship between different categorical variables and the difference in antibody MSI between hospitalized and non-hospitalized groups of patients. Spearman’s rank Correlation test was used to test the correlation between different antibody MSI and dilution factor for the neutralization assay. The general significance level was set at a p-value below 0.05. All analyses were performed using packages stats from the R statistical computing program v. 3.6.1 (Date of release 07/05/2019).

## 3. Results

### 3.1. Evolution of the IgG profile over time

Using the SirYus-CoViDiag® assay on 209 serum samples, the seropositivity stayed stable around 90 % for 6 months after an initial positive SARS-CoV-2 RT–PCR, before decreasing to 83.1 % between six and eight months (Fig. 2A).

**Figure 2.**
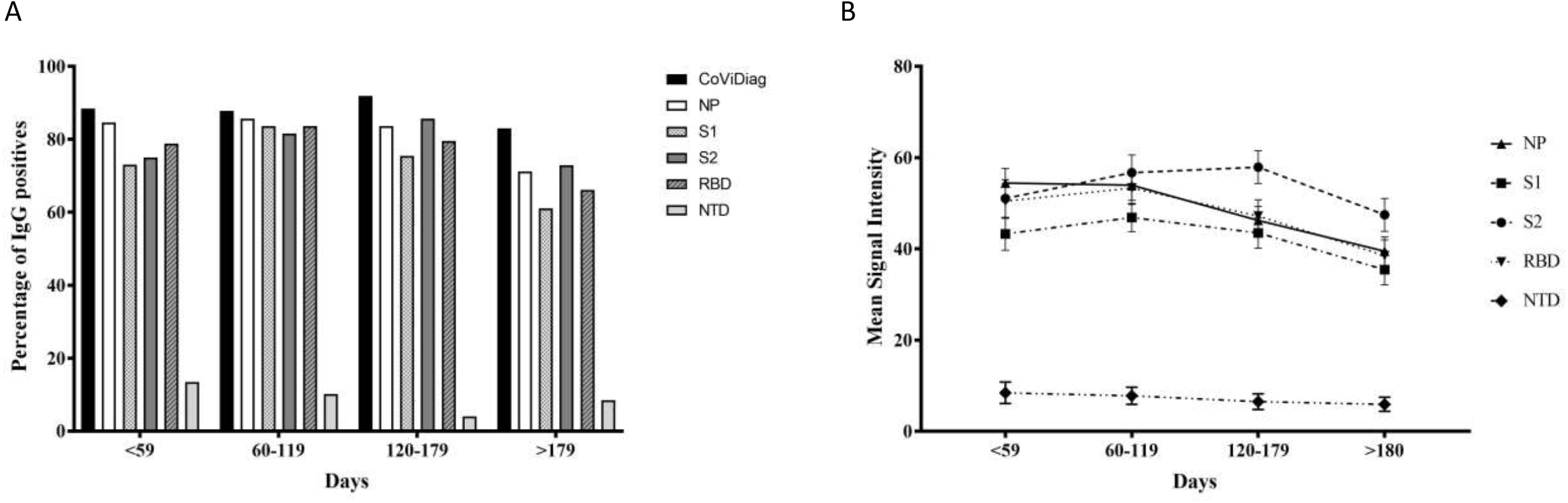
Evolution of the IgG profile over time. (A) Percentage of patients CoViDiag positive to anti-NP, anti-S1, anti-S2, anti-RBD, and anti-NTD IgG antibodies and (B) associated average IgG titers.

Positivities for each IgG considered individually are also reported based on the cut-offs set by the manufacturer. 54.1 % (n=113/209) samples were concomitantly positives for anti-NP, anti-S1, anti-S2 and anti-RBD antibodies and 9.1 % (n=19/209) for all 5 antibodies. 4.3 % (n=9/209) samples were positives for a single antibody (n=6 for anti-NP, n=2 for anti-S1 and n=1 for anti-S2) (Table 2). 80.9 % (n=169/209) samples were positives to anti-NP antibodies, allowing potential differentiation of infection from vaccination.

**Table 2.** Prevalence of the profile of IgG immune response. Percentage of positivity for antibodies against different antigens or combinaison of antigens.

|  | N | % Seroprevalence |
| --- | --- | --- |
| Samples | 209 |  |
| Single antibody |  |  |
| *NP* | 6 | 2.9 |
| *S1* | 2 | 1.0 |
| *S2* | 1 | 0.5 |
| *RBD* | 0 | 0 |
| *NTD* | 0 | 0 |
| *Total* | 9 | 4.3 |
| Combinaison of antibodies* |  |  |
| *NP+S2* | 5 | 2.4 |
| *NP+S1+S2* | 5 | 2.4 |
| *NP + S1 + RBD* | 6 | 2.9 |
| *NP+S2+RBD* | 13 | 6.2 |
| *S1+S2+RBD* | 6 | 2.9 |
| *NP+S1+S2+RBD* | 113 | 54.1 |
| *NP+S1+S2+RBD+NTD* | 19 | 9.1 |

\* For clarity purpose, only most common combinaisons with >2% seroprevalence are reported.

The kinetics of the IgG serum antibody response to individual antigens are presented in Fig. 2B. Average MSIs have been calculated for all samples depending on the time post RT-PCR to SARS-CoV-2. The anti-NP and anti-NTD antibody responses were the first to decrease, as their MSI started to decline after just two months (−0.9 % and -8.1 % between two and four months, respectively). The anti-S1 and anti-RBD response peaked after four months, before significatively decreasing over time (−7.8 % and -13 % between four and six months, respectively). The anti-S2 antibody response was the most delayed, with a peak level reached between four and six months. The different dynamics observed show the interest of detecting IgG response against multiple immunogenic domains to maintain elevated diagnostic sensitivity, especially long after infection.

### 3.2. IgG profile depending on the disease severity

Then we have investigated the ability for the multiplex assay to differentiate hospitalized (severe cases) versus non hospitalized (mild cases) patients, based on the first sample collected for each of the 61 patients in the early convalescent phase of the disease. For all five immunogenic domains, the MSI, corresponding to the levels of antibody are plotted in Fig. 3, depending on disease severity. For each given antigen, we have observed a trend of greater antibody response for hospitalized patients (MSI: NP= 56.5 ; S1=49.1 ; S2=59.4 ; RBD=54.8 ; NTD= 11.8; Average=46.3) compare to non-hospitalized ones (MSI: NP= 51.8 ; S1=37.4 ; S2=49.2 ; RBD=47.1 ; NTD=4.3 ; Average=37.9). However, the differences were not statistically different (p-value > 0.05, see supplementary Table 1).

**Figure 3.**
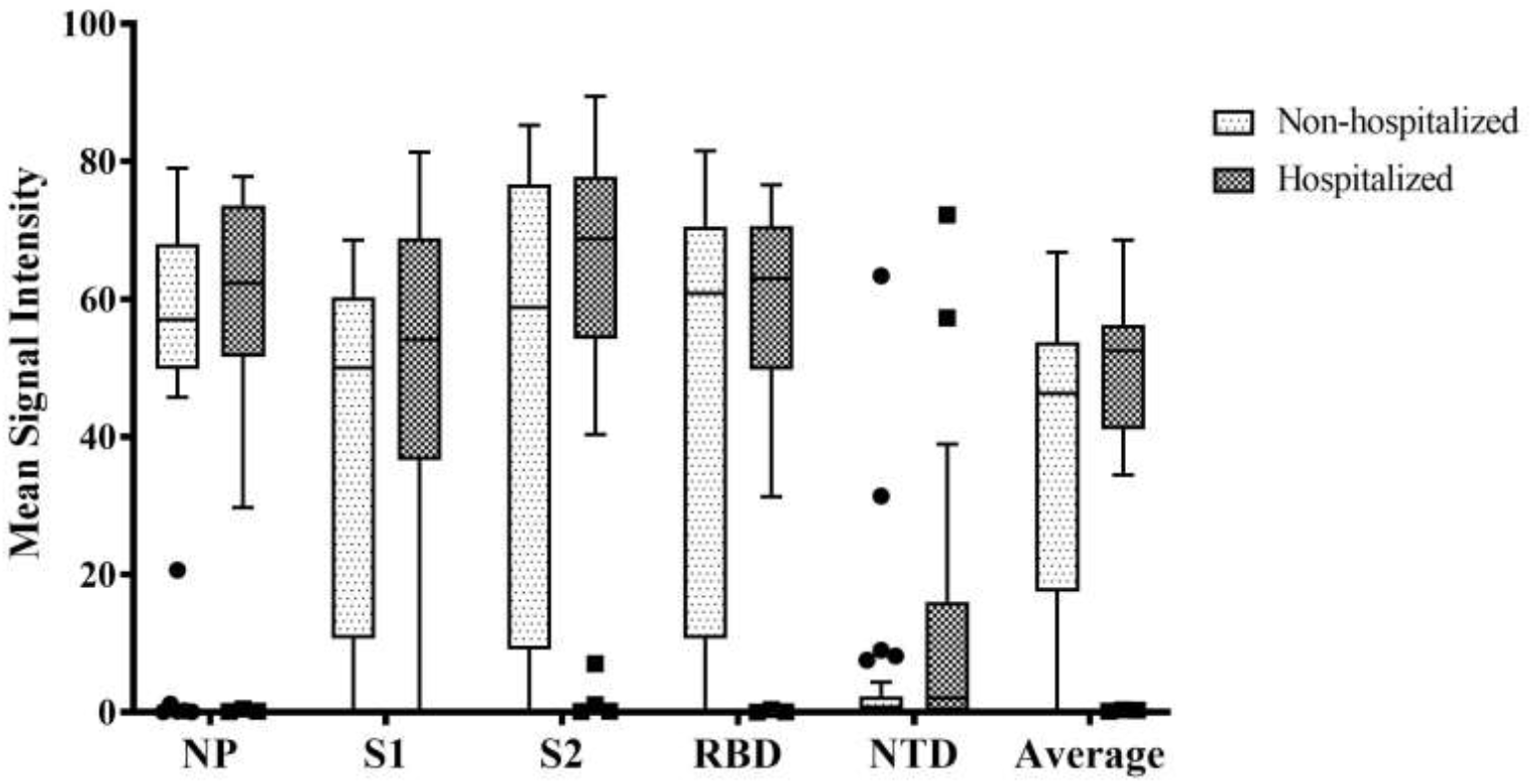
IgG profile depending on disease severity outcome. Distribution of the different IgG titers based on the MSI, considered individually, or altogether (average) for hospitalized (n=25) and non-hospitalized patients (n=34) just after infection.

### 3.3. Correlation between IgG profile and neutralizing antibody titers

Finally, we have evaluated the ability for the correlation between the different IgG levels response and the seroneutralization potential of the samples. For all five immunogenic domains, the mean intensity, corresponding to the levels of antibody response are plotted in Fig. 4 depending on the highest dilution of serum resulting in a 90 % decrease in infectivity. As expected, the best correlation (see supplementary Table 2) between individual IgGs and neutralizing antibody response was obtained for anti-RBD antibodies (r^2^=0.72). The correlation was very similar between anti-S1 (r^2^=0.67) and anti-S2 (r^2^=0.66) antibodies. However Anti-NP (r^2^=0.59) and anti-NTD (r^2^=0.47) antibodies titers were less correlated with the neutralizing antibody titers. Interestingly, the combination of the 5 different antibody responses, allowed to slightly increase the correlation to (r^2^=0.74).

**Figure 4.**
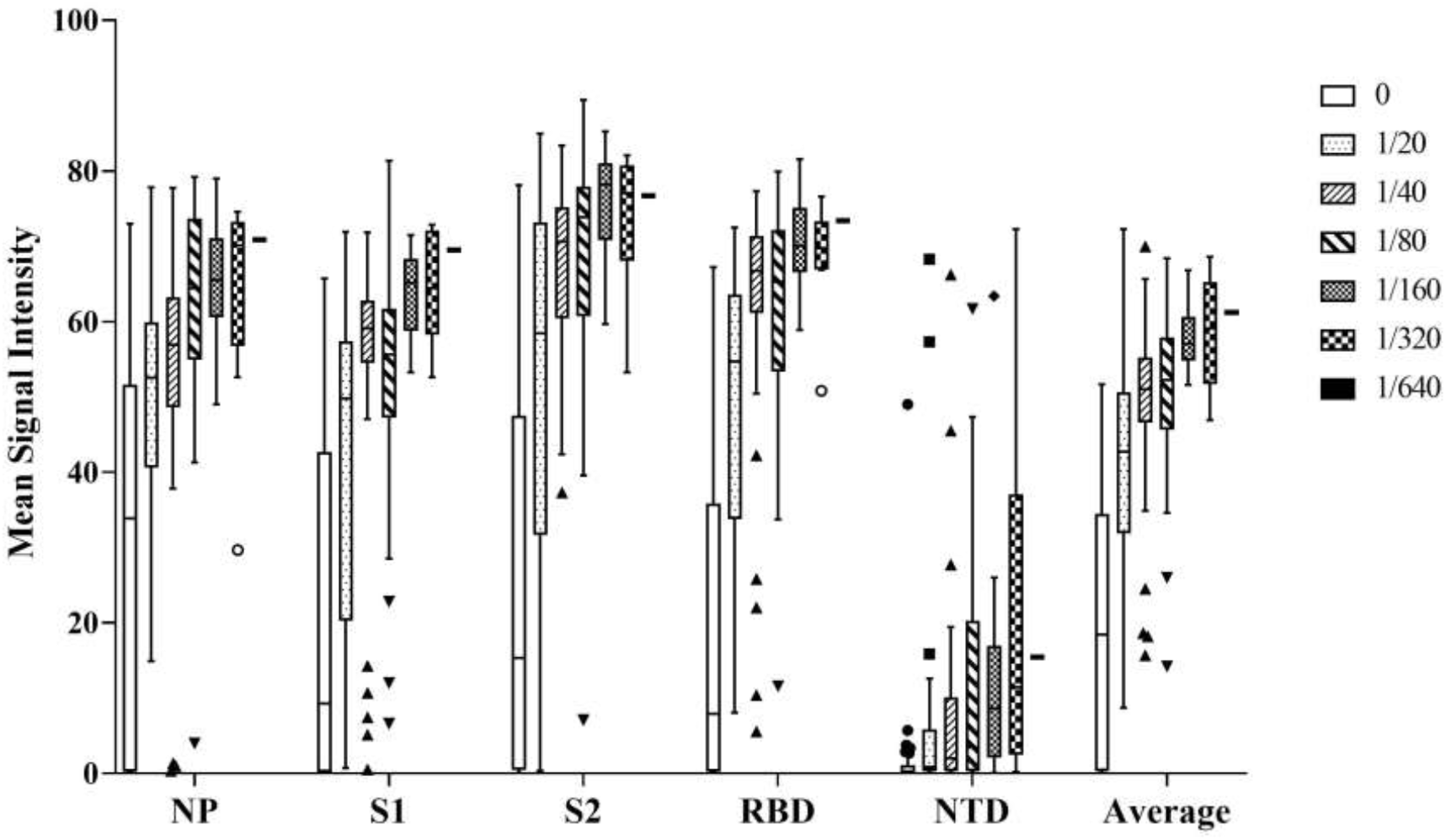
Correlation of the different IgG titers with serum neutralization titers. IgGs titers are based on the MSI considered individually, or altogether (average). Neutralizing antibody titers are based on the serum dilution factor to neutralize 90 % of infected cells.

## 4. Discussion

In a previous study based on the same set of sample, we have found equivalent to improved diagnostic performances, especially for ancient infections, for the CoViDiag® multiplex IgG assay compare to other IgG commercial serological assays (13). Is is now generally admitted that antibody levels are weaker for asymptomatic and mild form of the disease and can decrease over time (8). Hence in the present work, we have investigated the detailed profile of the IgG immune response over an eight months period in a multiplex assay, using samples of hospitalized and non-hospitalized patients. Then we have comparde the results with neutralizing antibody levels.

We have observed that most patients develop a global immune response against multiple immunogenic domains. Even over a 8 months period, more than a half of the samples were positives to anti-NP, anti-S1, anti-S2, and anti-RBD antibodies, concomitantly. Those result confirm the possibility to develop serological assays based on different antigens. Anti-NTD antibodies are more scarce but most patients SARS-CoV-2 infected develop antibodies against the NP antigen allowing potential differentiation of infection from vaccination.

As expected, the different IgGs titers decreased over time, but with different dynamics. Elevated levels of anti-S2 seem to last longer. Therefore the detection of anti-S2 antibodies may be of interest to maintain elevated diagnostic sensitivity longer after infection. The evolution of anti-S1 and anti-RBD titers is very similar, as RBD constitutes a domain of the Spike 1 protein.

For all the tested IgGs, we have found higher titers for hospitalized patients than for non-hospitalized ones. However, the differences were not statistically significant as a large number of patients had no immune response detected for individual antigens, independently of the disease severity.

It is noteworthy that most commercial assays performances have been established at the beginning of the epidemic, when samples from hospitalized patients were the easier to collect. For people presenting a weaker immune response, multiplexing allows to test for extra domains that may help to slightly increase diagnostic sensitivity without compromising for diagnostic specificity.

Except for anti-NTD antibodies, all different IgGs titers were positively correlated with the neutralizing antibody titers. This result is not surprising considering our previous observation showing that anti-NP, anti-S1, anti-S2, and anti-RBD antibodies are concomitantly present in patient’s sera. As expected, the best correlation for individual antigen is obtained for antibodies targeting the virus RBD domain which is known to be involved in the penetration of the cells by the virus. However the average combination of all five antigens slightly increased the correlation, strengthening the interest for multiplexing.

## 5. Conclusion

Beyond the diagnosis of SARS-COV-2 infection, tools delivering a global picture of the patients’ immune response may also be of interest to improve the management and care of the patients and populations. Our results show that elevated IgGs titers against multiple viral epitope may be more characteristic of symptomatic patients, and correlates well with neutralizing antibodies. We recommend using assays targeting IgGs for the evaluation of a long lasting population protection and collective immunity. Furthermore, multiplexed assays have the potential to slightly increase diagnostic performances, especially for ancient or weak infections and be more representative of immune protection. For future epidemical studies, as the vaccination based on the Spike protein progresses, multiplex serological assays may also help to differentiate vaccination from viral infection.

## Data Availability

Raw data and results are available in supplementary data.

## Funding

**Laboratory’s own resources**

## Declaration of competing interest

Authors Rémi Malbec, Gaël Even and Christophe Audebert are employees of GD Biotech, while Pauline Ponthieu, Pauline Follet, Vianney Souplet and Christophe Olivier are employees of Innobiochips, providing the CoViDiag® assay kits for this study.

## Supporting information captions

**Supplementary Table 1**. Mean Signal Intensity (MSI) of the antibody response to different antigens for hospitalized and non-hospitalized patients.

**Supplementary Table 2**. Correlation between individual antigen titers and neutralizing antibody titers.

## Notes

### Competing Interest Statement

Authors Remi Malbec, Gael Even and Christophe Audebert are employees of GD Biotech, while Pauline Ponthieu, Pauline Follet, Vianney Souplet and Christophe Olivier are employees of Innobiochips, providing the CoViDiag assay kits for this study.

### Funding Statement

Laboratory's own ressource

### Author Declarations

The study was approved by the institutional review board of the Amiens University Medical Center (number PI2020_843_0046, 21 April 2020).

