## Supplemental Table 1 for "A multiplex serological assay for the characterization of IgG immune response to SARS-CoV-2"

### Slide 1
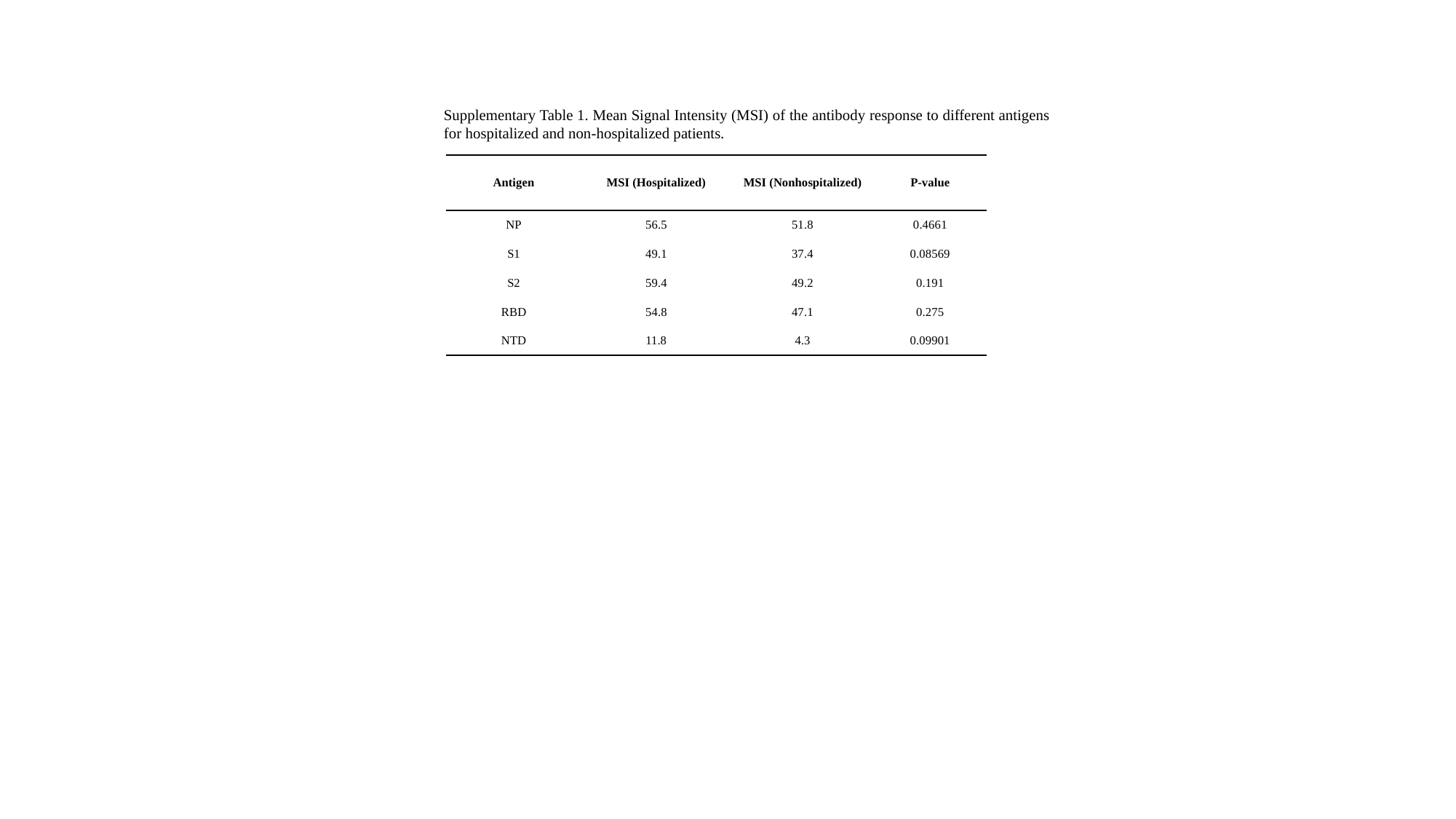

Supplementary Table 1. Mean Signal Intensity (MSI) of the antibody response to different antigens for hospitalized and non-hospitalized patients.
| Antigen | MSI (Hospitalized) | MSI (Nonhospitalized) | P-value |
| --- | --- | --- | --- |
| NP | 56.5 | 51.8 | 0.4661 |
| S1 | 49.1 | 37.4 | 0.08569 |
| S2 | 59.4 | 49.2 | 0.191 |
| RBD | 54.8 | 47.1 | 0.275 |
| NTD | 11.8 | 4.3 | 0.09901 |
