## Supplemental Table 2 for "A multiplex serological assay for the characterization of IgG immune response to SARS-CoV-2"

### Slide 1
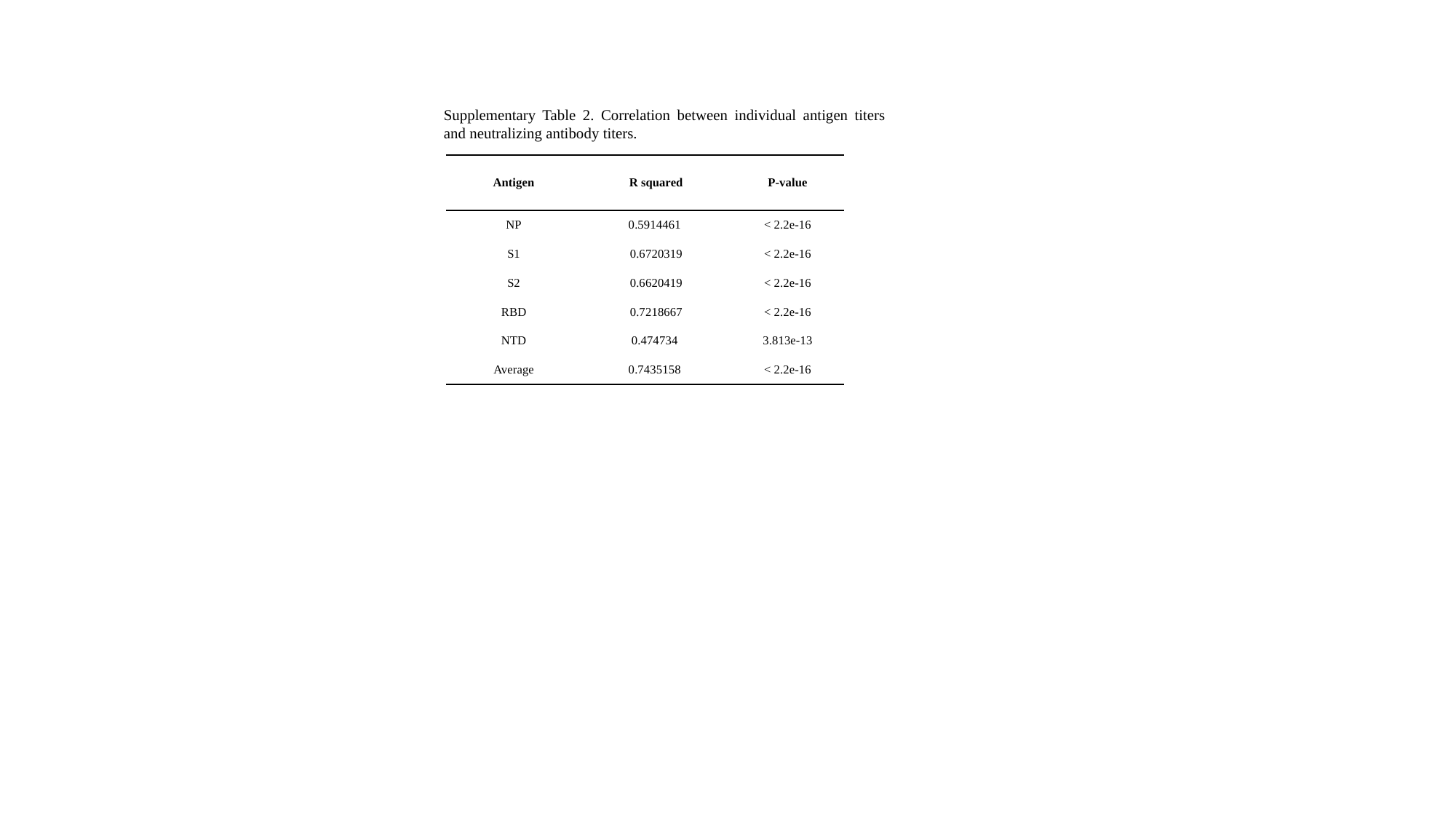

Supplementary Table 2. Correlation between individual antigen titers and neutralizing antibody titers.
| Antigen | R squared | P-value |
| --- | --- | --- |
| NP | 0.5914461 | < 2.2e-16 |
| S1 | 0.6720319 | < 2.2e-16 |
| S2 | 0.6620419 | < 2.2e-16 |
| RBD | 0.7218667 | < 2.2e-16 |
| NTD | 0.474734 | 3.813e-13 |
| Average | 0.7435158 | < 2.2e-16 |
